# Heterogeneity in COVID-19 Pandemic-Induced Lifestyle Stressors and Predicts Future Mental Health in Adults and Children in the US and UK

**DOI:** 10.1101/2021.08.10.21261860

**Authors:** Aki Nikolaidis, Jacob DeRosa, Mirelle Kass, Irene Droney, Lindsay Alexander, Adriana Di Martino, Evelyn Bromet, Kathleen Merikangas, Michael Peter Milham, Diana Paksarian

## Abstract

Identifying predictors of mental health symptoms after the initial phase of the pandemic may inform the development of targeted interventions to reduce its negative long-term mental health consequences. In the current study, we aimed to simultaneously evaluate the prospective influence of life change stress, personal COVID-19 impact, prior mental health, worry about COVID-19, state-level indicators of pandemic threat, and socio-demographic factors on mood and anxiety symptoms in November 2020 among adults and children in the US and UK. We used a longitudinal cohort study using the Coronavirus Health Impact Survey (CRISIS) collected at 3 time points: an initial assessment in April 2020 (“April”), a reassessment 3 weeks later (“May”), and a 7-month follow-up in November 2020 (“November”). Online surveys were collected in the United States and United Kingdom by Prolific Academic, a survey recruitment service, with a final sample of 859 Adults and 780 children (collected via parent report). We found subtypes of pandemic-related life change stress in social and economic domains derived through Louvain Community Detection. We assessed recalled mood and perceived mental health prior to the pandemic; worries about COVID-19; personal and family impacts of COVID-19; and socio-demographic characteristics. Levels of mood symptoms in November 2020 measured with the circumplex model of affect. We found 3 life change stress subtypes among adults and children: Lower Social/Lower Economic (adults and children), Higher Social/Higher Economic (adults and children), Lower Social/Higher Economic (adults), and Intermediate Social/Lower Economic (children). Overall, mood symptoms decreased between April and November 2020, but shifting from lower to higher-stress subtypes between time points was associated with increasing symptoms. For both adults and children, the most informative predictors of mood symptoms in November identified by conditional random forest models were prior mood and perceived mental health, worries about COVID, and sources of life change. The relative importance of these predictors was the most prominent difference in findings between adults and children, with lifestyle changes stress regarding friendships being more predictive of mood outcomes than worries about COVID in children. In the US, objective state-level indicators of COVID-19 threat were less predictive of November mood than these other predictors. We found that in addition to the well-established influences of prior mood and worry, heterogeneous subtypes of pandemic-related stress were differentially associated with mood after the initial phase of the pandemic. Greater research on diverse patterns of pandemic experience may elucidate modifiable targets for treatment and prevention.

## 1. Introduction

As the COVID-19 pandemic has led to worldwide illness, death, and disruptions in daily life, its effects on emotional well-being have become a public health priority.^1^ Overall, studies have demonstrated high levels of anxiety and related conditions in samples of adults,^1,2^ with fewer investigations in children and adolescents.^3^ Most studies have been cross-sectional, and many have focused on at risk subgroups (e.g., healthcare and essential workers).^4^ Longitudinal studies of general population samples have found that mental health problems initially increased compared to pre-pandemic levels,^5–7^ followed by a decrease as the pandemic continued.^8–10^ This is consistent with prior research on population mental health responses to disaster, which has shown that while transient increases in symptomatology are common, long-lasting problems are rare.^11^ Longitudinal studies that identify predictors of poor mental health versus resilience after the initial phase of the pandemic could help to inform the development of targeted interventions to reduce long-term mental health consequences.

Prior disaster mental health research has consistently found that pre-disaster mental health and the degree of fear and worry during a disaster are primary determinants of mental health outcomes.^12^ Mental health responses to disaster are also worse among those who are directly and severely impacted, and vary by socio-demographic characteristics such as younger age, gender, lower income, and minority race/ethnicity.^13^ Consistent with these findings, initial work by our team and others highlighted prior mental health and the degree of worry about COVID-19 as key predictors of mood in the initial months of the COVID-19 pandemic.^2,14^ Along with others, our prior work also highlighted the importance of the widespread changes in daily life and accompanying stressors induced by the pandemic for mental health.^9,15–20^ Together these prior findings underscore the multifactorial influences of the pandemic among families and individuals. A greater understanding of the long-term role of pandemic-related life change stress, in relation to other risk factors for poor mental health, is needed in order to understand potential avenues for intervention.

The goal of the current study was to extend our previous findings by using a longitudinal design to characterize the role of life change stress in shaping mood after the initial phase of the COVID-19 pandemic. After an initial assessment in April 2020 (“April”), we reassessed participants three weeks later (“May”) to capture rapid changes during the beginning of the pandemic, and then conducted a 7-month follow-up in November 2020 (“November”) among our cross-national sample of adults and children (assessed via self and parent report, respectively) in the US and UK. Our aims were to: 1) Define subtypes of life change stress at 3 weeks and their associations with mood and other correlates. 2) Assess the stability of subtype membership across time points, and how subtype transitions were associated with changes in mood and worries about COVID-19. 3) Identify the importance of life change stress subtypes in predicting mood at follow up, while considering other factors including demographics, recalled prior mood and mental health, worries about COVID-19, personal and family pandemic impact, and objective state-level indicators of pandemic threat. To our knowledge, this is the first longitudinal study to simultaneously investigate the roles of life change stresses, personal COVID-19 impact, prior mental health, worry about COVID-19, state-level indicators, and socio-demographic factors in levels of mood among adults and children in multiple countries.

## 2. Methods

### 2.1 Samples

Data were collected through Prolific Academic (PA; https://www.prolific.ac/), an online crowdsourced survey recruitment service (Table 1). Participants who signed up to join the PA participant pool received monetary compensation for their time. PA offers samples that broadly reflect the population distributions of age, sex, and race in the US and UK; participants have been shown to be more diverse and provide higher quality data than similar data collection platforms.^21^ In March 2020 we requested four samples of 1,500 participants from the US and UK, both adult self-report and parent report. Portions of the sample were targeted at regions that were more severely impacted by COVID-19 in late March 2020 (New York, California, London, and Manchester). For the child (parent report) sample, users were screened based on having a child between 5 and 18 years old and were asked to report on their oldest child in that age range; reports received for children over age 25 were excluded. No additional exclusion criteria were given to PA.

**Table 1:**
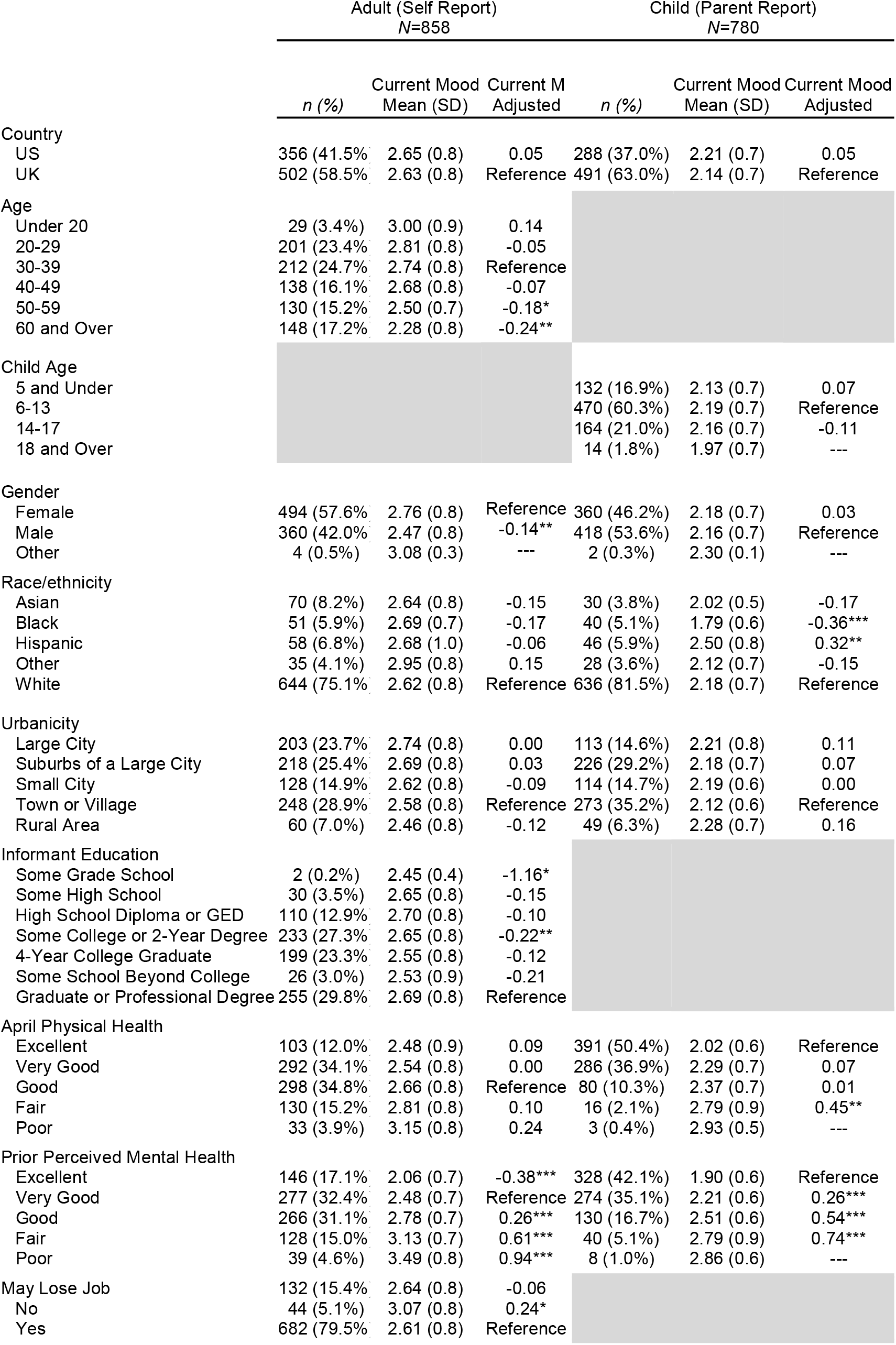
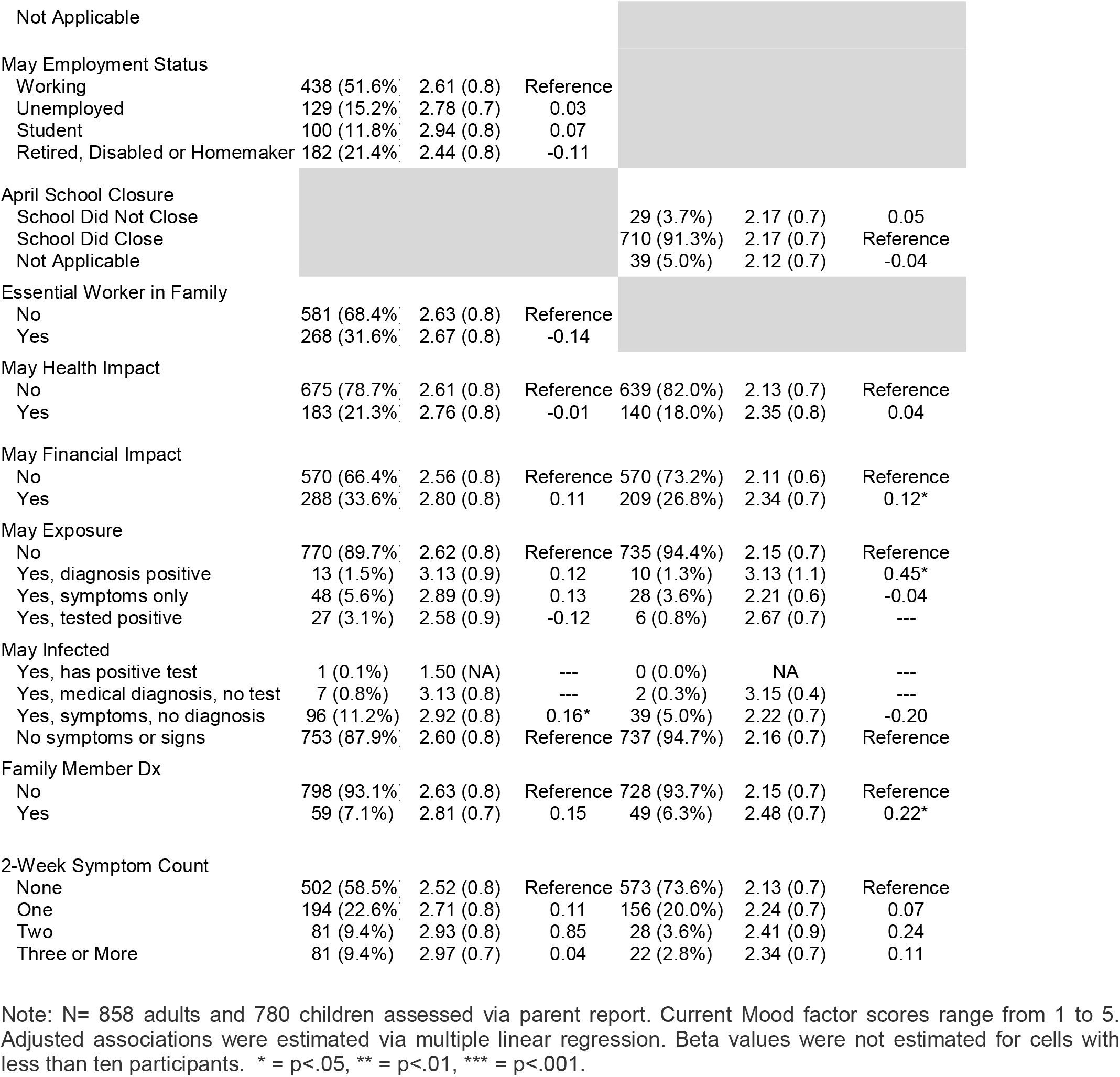
Sample characteristics with mean Current Mood scores and adjusted associations with Current Mood.

Three waves of data were collected in 2020: at baseline (April 7^th^-17^th^), 3 weeks (April 24^th^-May 4^th^), and 7-month follow up (October 31^st^-November 23^rd^). For the present analyses we excluded participants with uncompleted forms and incomplete information for subtyping (described below). Final analytic sample sizes were: 3,259 in April (1,793 adults and 1,466 parents), 2,553 at 3 weeks (1,380 adults and 1,173 parents), and 1,639 in November (859 adults and 780 parents) (see eTable 1).

Exemption from IRB oversight was approved by the Advarra Institutional Review Board. PA participants were required to agree to PA’s Terms of Service notification (https://prolific.ac/assets/docs/Participant_Terms.pdf) to complete surveys. Per the IRB exemption, no additional informed consent was required. Comparisons between the demographics of the included sample and both the drop out and Census for US and UK are included in the supplement (eTable 2, eTable 3).

### 2.2. Measures

The Coronavirus Health Impact Survey (www.crisissurvey.org) was administered online via REDCap software at all 3 time points. Abbreviated baseline versions of CRISIS v0.3 were administered in April and follow-up versions of CRISIS v0.3 were administered in May. The CRISIS v0.3 baseline forms were revised and expanded for the November follow-up (available by request).

#### 2.2.1. Mood

Ten items from the circumplex model of affect^22,23^ (see Supplement) rated on a 5-point scale were included to measure mood. Prior mood was assessed at baseline by asking participants to recall the 3 months prior to the pandemic. Mood over the past 2 weeks (“Current mood”) was included from the May and November time points. Based on excellent unidimensional fit (see eTable 4), items were averaged to generate a total score (range: 1 to 5), with higher scores indicating worse mood. Unadjusted associations with mood are included in the supplement (eTable 5, eTable 6).

#### 2.2.2. Predictors

##### Participant characteristics

Age, sex, self- or parent-rated mental and physical health, urbanicity, informant education, and employment (adults only) were reported on the CRISIS in April. Race was reported to PA and combined with CRISIS information on Hispanic ethnicity to generate the following categories: Hispanic, non-Hispanic White, non-Hispanic Black, Asian, and Other race/ethnicity. For children, parent race was used as a proxy for child race. Perceived mental health prior to the pandemic, and physical health during the pandemic, were rated on 5-point scales from “excellent” to “poor.”

##### Personal COVID-19 impact

Perceived exposure to SARS-CoV-2, COVID-19 symptoms, family member diagnosis of COVID-19, and impacts on family members such as hospitalization, quarantine, and job loss because of COVID-19 were reported on the CRISIS in May with respect to the past 2 weeks. In April, participants reported whether they or a household member were an essential worker.

##### COVID-19 Worries in the past two weeks

In May, participants reported on a five-point Likert scale how worried they have been during the past two weeks about infection, friends and family being infected, and possible impacts on physical and mental health, as well as time spent reading or talking about COVID-19 and hope that the pandemic will end soon. Based on psychometric analyses (described in supplement), these were averaged to generate a total score (range: 1 to 5; hereafter referred to as “COVID-19 Worries”).

##### Life Changes due to the pandemic in the past two weeks

Downstream and subjective impacts of structural changes, such as changes in social contacts, effects on family relationships, changes in living situation, food insecurity, and stressors associated with these changes were reported on the CRISIS (14 items; see Supplement). Life change stress subtypes were derived independently at all 3 timepoints based on these items (described below).

##### Objective Government Response Tracker (OxCGRT) Indices

Following the model of Vibert et al., (in prep), state-level indices from OxCGRT^24^ were used to indicate objective levels of COVID-19 threat in each US participant’s state. Specifically, we included: Containment and Health Index (lockdown restrictions and closures related to COVID-19), Economic Support Index (economic relief for households provided by the government), Confirmed Cases by state per 1,000,000 residents, and Confirmed Deaths by state per 1,000,000 residents (see Supplemental Methods). These indices covered the time period: January 1st, 2020 to March 19th, 2021. Average values of each index were calculated from April 1st to May 31st. Regionally specific containment data were not available for the UK, and so our analyses using containment included only US data.

### 2.3 Analysis

#### Community Detection Based Subtyping (Aims 1 & 2)

Life change stress subtypes were derived using Louvain community detection (Aim 1; LCD; Figure 1; eFigure 1; see supplemental methods), enhanced through bootstrap aggregation (i.e., bagging), as in our prior study.^20^ Structural similarity across samples and timepoints was confirmed using Pearson’s correlation. LCD is a clustering technique that selects clusters to maximize within-cluster coherence and between-cluster segregation. Subtype differences between individual items are described in the supplement (eTable 7). We compared demographic characteristics and COVID-19 impact indicators across May subtypes using one-way ANOVA (Table 2; eTable 8). We also estimated mean mood and COVID-19 Worries scores according to patterns of change in subtype membership across the 3 timepoints and used t-tests to test mean differences (Aim 2; Figure 2).

**Figure 1.**
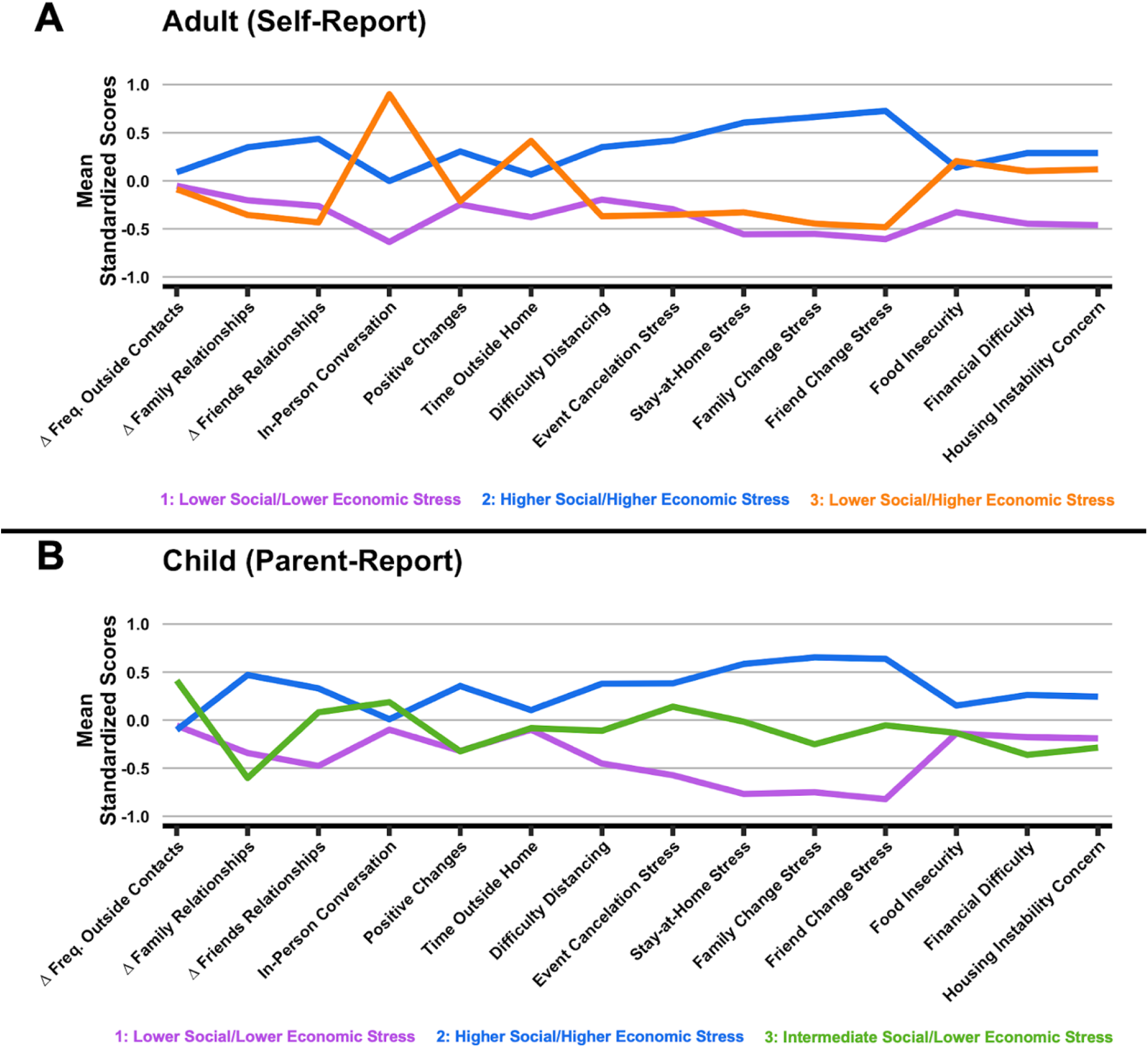
May Life Change Stress Subtypes. Note: A. Life Change Stress Subtype profiles for adults (top) and children (bottom) in May 2020. Mean normalized profile loadings are displayed on the y-axis. Δ Family Relationships and Δ Friends Relationships are coded so that higher scores indicate worsening quality of relationships. In-Person Conversation, Positive Changes, and Time Outside Home are coded so that higher scores indicate less conversations, positive changes, and time spent outside. Adult Report: Purple (1): Lower Social/Lower Economic Stress, Blue (2): Higher Social/Higher Economic Stress, Orange (3): Lower Social/Higher Economic Stress. Parent Report: Purple (1): Lower Social/Lower Economic Stress, Blue (2): Higher Social/Higher Economic Stress, Green (3): Intermediate Social/Lower Economic Stress.

**Table 2:**
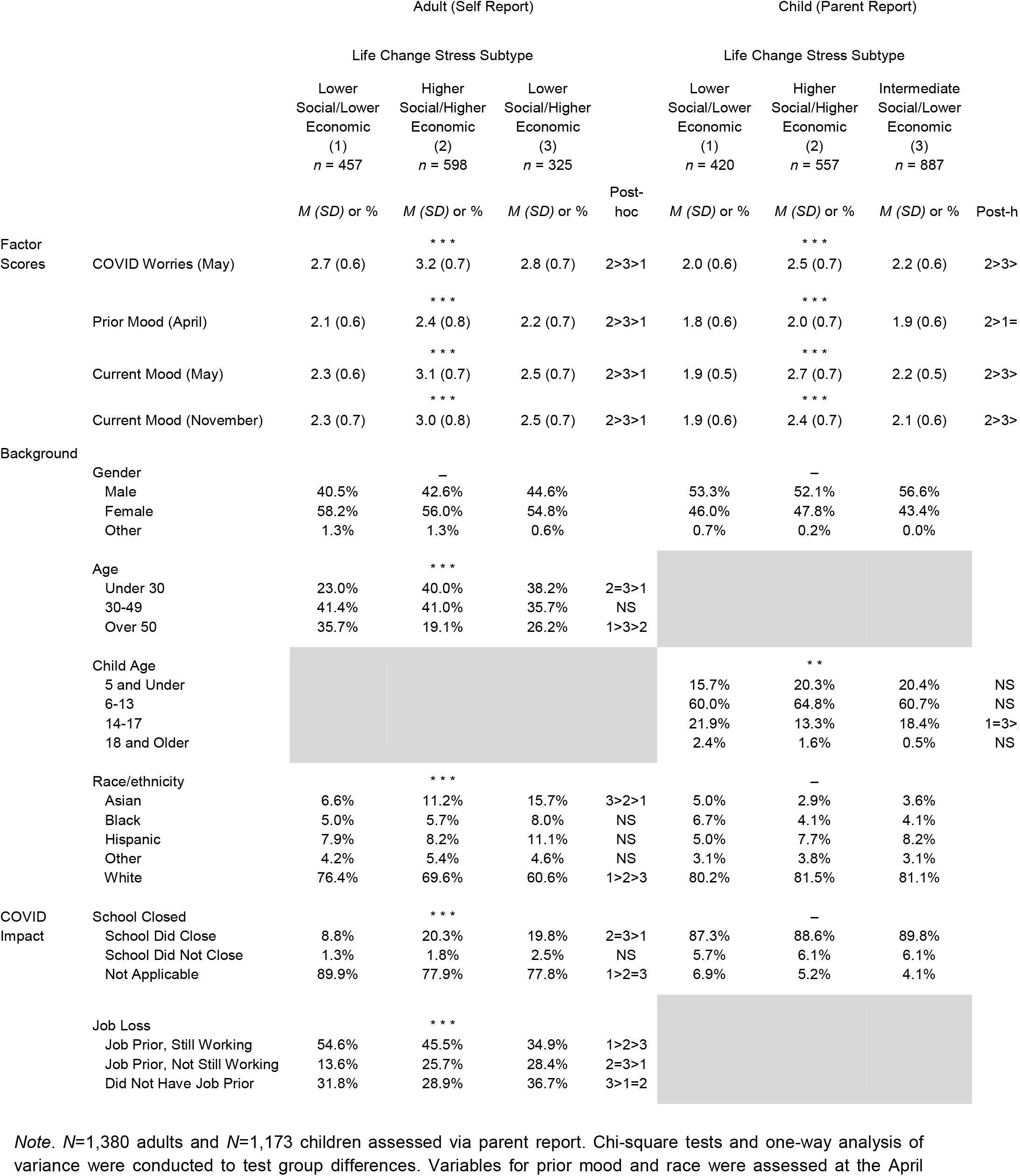
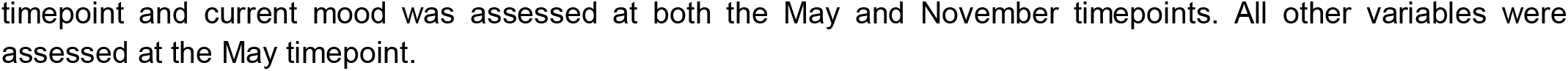
Mood, COVID Worries, demographic characteristics, and COVID impact indicators across May life change stress subtypes.

**Figure 2:**
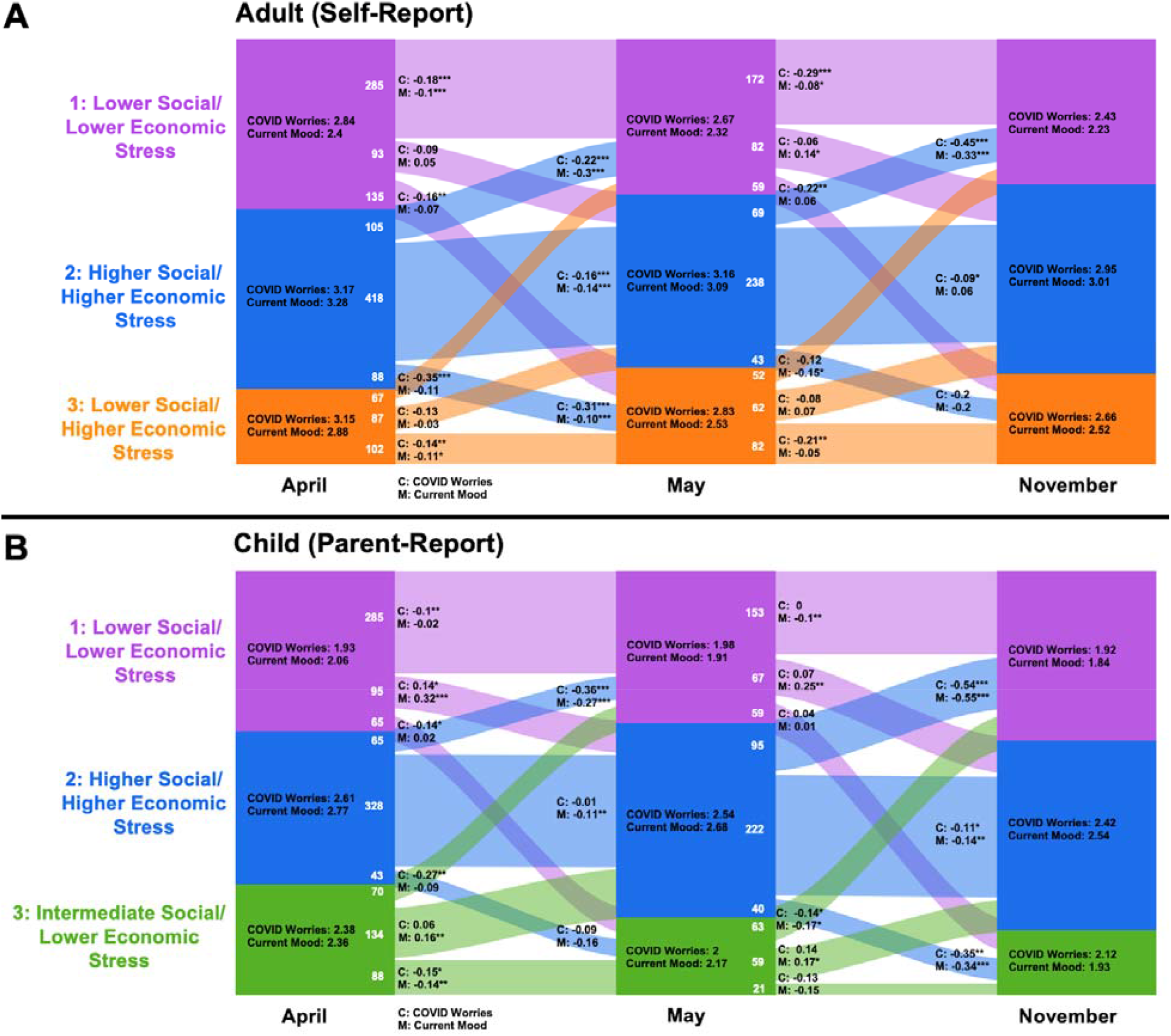
Stability and change in Life Change Stress Subtype across time points with mean mood and COVID Worries scores by group. Note: Colored areas correspond to the proportion of participants in each subgroup at each time point. The numbers of individuals moving between Subtypes are given in white, with proportions indicated by the width of the colored paths. Mean COVID Worries and Current Mood scores are represented by Subtype in the center of each column. COVID Worries (C:) and Mood (M:) are represented in the change paths for each change group. T-tests compared the change in the scores between time points for each change group: ^*^ = p < 0.05, ^**^ = p < 0.01, ^***^ = p < 0.001.

#### Conditional Random Forest (Aim 3)

Finally, we used conditional random forest (CRF) to predict mood in November based on prior mood and perceived mental and physical health, COVID-19 worries factor scores, May life change stress subtype, OxCGRT indices, socio-demographic characteristics, and personal and family COVID-19 impact. Random forest (RF) is a robust predictive machine learning technique known for its ability to handle dependencies and interactions between predictor variables (See Supplemental Methods).^26,27^ CRF overcomes some limitations of traditional RF and estimates how much each variable contributes to overall model accuracy, conditioned on other included variables.^28^ We built 3 sets of CRF models: a base model including all predictors described above, using life change stress subtypes; a second model which replaced life change stress subtypes with their constituent items; and a third model in the US sample alone which added the OxCGRT indices.

Because our prior study demonstrated highly reproducible findings across the US and UK, and to maximize power for prediction modeling, analyses were conducted combining the US and UK samples. The proportion of missing data in the analytic sample was less than 2% for all variables and missing data were handled via model-wise deletion.

## 3. Results

### 3.1 Sample Characteristics

Table 1 shows demographic characteristics and COVID-19 impact indicators for the November 2020 sample, along with means and standard deviations of November mood scores. The majority of adults were females (57.6%), white (75.1%), and aged 30-39 (24.7%), while the majority of children were males (53.6%), white (81.5%), and aged 6-13 years (60.3%). Overall, mood symptom and COVID-19 Worries levels decreased between the April and November both adults (mood: -0.13; p<.0001; COVID-19 Worries: -0.35; p<.0001) and children (mood: -0.17; p<.0001; COVID-19 Worries: -0.16; p<.0001). Differences in November mood by participant characteristics are displayed in Table 1. Mean COVID-19 Worries and prior mood scores by demographics and COVID-19 impact indicators are presented in eTables 5 and 6.

### 3.2. Life change stress subtypes (Aim 1)

Life change subtypes derived from the 3-week time point (Figure 1) were named based on relative ratings of social/interpersonal and economic stressors (See eTable 7 and supplemental results). Adult subtypes were: *Lower Social/Lower Economic stress* (purple; 1), *Higher Social/Higher Economic stress* (blue; 2), and *Lower Social/Higher Economic stre*ss (orange; 3). Among children, subtypes were: *Lower Social/Lower Economic* (purple; 1), *Higher Social/Higher Economic* (blue; 2), and *Intermediate Social/Lower Economic stress* subtype (orange; 3).

### 3.3. Longitudinal Subtype Change (Aim 2)

Figure 2 shows the stability of, and change in, subtype membership across time points, with accompanying mean scores for COVID-19 Worries and mood. Most participants stayed in the same subtype across time points. Specifically, 805/1,380 adults and 747/1,173 children remained stable between April and May, and 492/859 adults and 396/780 children remained stable between May and November. Subtype 3 (Lower Social/Higher Economic for adults and Intermediate Social/Lower Economic for children) appeared the least stable. We saw an overall trend of either decrease or no change in COVID-19 Worries and mood across timepoints and subtypes for both adults and children. The only significant increase among adults was in mood for those who moved from the Lower Social/Lower Economic subtype in May to the Higher Social/Higher Economic subtype in November (estimate=0.14, p<.05). More increases were seen among children. Children who changed from Lower Social/Lower Economic to Higher Social/Higher Economic showed increases in COVID Worries (April-May only; estimate=0.14, p<.05) and mood (April-May: estimate=0.32, p<.001; May-November: estimate=0.25, p<.01). Children who moved from Intermediate Social/Lower Economic to Higher Social/Higher Economic showed increases in mood symptoms across both transitions (April-May: estimate=0.16, p<.01; May-November: estimate=0.17, p<.05).

### 3.4. Predicting November Mood (Aim 3)

Conditional random forests including participant characteristics, COVID-19 impact indicators, prior mood and mental health, COVID-19 worries, and life change subtypes predicted substantial variation in November mood (Figure 3; Adult R^2^=59.4%; Child R^2^=47.8%). The most important predictors were prior mood, COVID-19 Worries, prior perceived mental health, and life change subtype for adults and children. While COVID-19 Worries was the second most informative predictor for adults, life change subtypes were the second most informative for children. When life change stress was included as individual items, social/interpersonal stress items showed the highest conditional variable importance for both adults and children, and the variance explained increased (Figure 3; Adult R^2^+5.4%; Child R^2^+8.6%). For the US samples of adults and children, adding indicators of US state-level COVID-19 threat from OxCGRT (eFigure 2) modestly increased variance explained, but these variables exhibited lower conditional importance (Adult R^2^+3.1%; Child R^2^+1.1%) relative to other variables.

**Figure 3:**
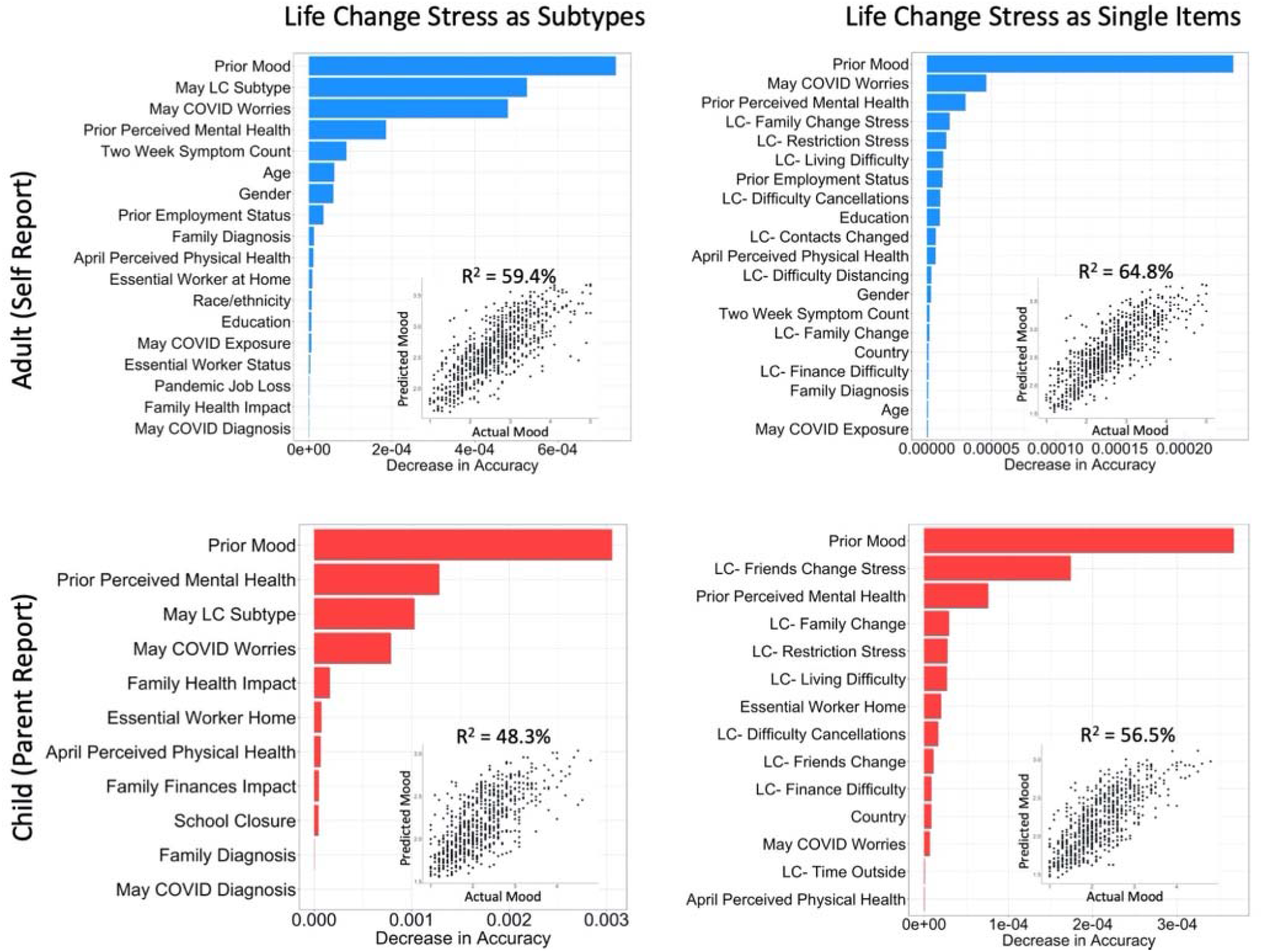
Results of conditional random forest models predicting Current Mood in November 2020. Note: Conditional Random Forest Models in adults (top; n=827) and children (bottom; n=750) showing the conditional variable importance of each predictor. Variable importance is calculated based on decreases in accuracy when a variable is removed conditioned on all other variables in a model using balanced bootstrapped partitions of the data to ensure unbiased importance estimates. Only variables with importance values greater than zero are shown. Life change stress was included as data-derived subtypes on the left and as individual indicators on the right.

## 4. Discussion

In a longitudinal study covering the beginning of the COVID-19 pandemic and a six month follow-up between April and November 2020, we found that prior mood, worry about COVID-19, and profiles of pandemic-induced life change stress were the most important predictors of mood symptom levels in November 2020. The importance of prior mood and worry about COVID-19 is supported by extensive prior studies of mental health during the pandemic,^15,17,29,30^ as well as studies following other large scale traumatic events (i.e., natural disasters, terrorist attacks, nuclear reactor meltdowns).^12,31–33^ Of note, the relative importance of these three predictors was the most prominent difference in findings between adults and children, with lifestyle changes stress (particularly that related to friendships) being more predictive of mood outcomes than worries about COVID-19 in children. Country did not emerge as an important predictor of later mood, implying similarity of predictors across sites. The importance of the life change stress subtypes underscores the heterogeneity in the effects of the pandemic on people’s lives.

In the current study, patterns of life change stress were highly reproducible across samples and over time. They largely captured individual variation in secondary effects of the pandemic beyond medical illness, such as changes in social relationships and financial security. Their importance is consistent with other studies of the COVID-19 pandemic, which have found associations of social distancing, stay-at-home orders, and financial strain with mental health.^34–36^ Our analysis of stability and switching life change stress subtypes indicated that for most groups, mood symptoms decreased between time points. This is consistent with other COVID-19 research showing that mental health symptoms rose at the start of the pandemic and subsequently declined.^9,16^ Higher mood levels in November may be an indicator of risk for prolonged mental health problems following the pandemic, although this needs to be confirmed with additional follow-up. Groups who deviated from the decreasing pattern were those who changed into a high-stress subtype between study waves, further demonstrating the importance of variation in life change stress as a contributor to mental health. The greater importance of life change stress subtypes over objective state-level indicators of pandemic threat may be because they are more proximally tied to mental health.

Although we cannot infer causal relationships, our longitudinal results may help to inform the targeting of intervention strategies and anticipate health services needs during future crises.^38^ Individuals with higher levels of symptoms prior to the pandemic are at risk of continued poor mental health during the pandemic, and should be regarded as a vulnerable subgroup.^15,17,39^ Research from prior disasters and the current pandemic has demonstrated the importance of reliable information in shaping people’s degree of fear and worry about an event.^40,41^ Therefore, national-level efforts to provide more high-quality information about COVID-19 and increase trust in legitimate information sources may help to decrease mood-anxiety symptoms among the general public. Economic stressors captured in the life change stress profiles may be addressed through policy interventions that decrease housing instability, food insecurity, and economic hardship.^42,43^ Sources of social and interpersonal stress may vary widely and therefore be more difficult to intervene on from a population level. However, strategies that may be useful include the provision of psychoeducational and coping resources,^44^ implementation of programs to reduce domestic and intimate partner violence and provision of resources for those affected,^45^ and increasing availability of formal and informal mental health services through telemedicine and other alternative venues.^46,47^

The major limitation of this study is the use of a web-based convenience sample, which raises concerns about the potential for selection biases and may limit the generalizability of our findings. For example, if selection into the sample differed by both mental health and life change stress indicators, this could have inflated the association between life change stress and mood. Use of this sample was motivated by the need to quickly develop and deploy the CRISIS questionnaire. CRISIS being used in several studies across the world will allow for the comparison of results across sample types and locations. In addition, we relied on recall for measurement of pre-pandemic mental health, and relied on parent reports for children, which may be less accurate for older children and for internalizing symptoms across ages.^48^ Furthermore, our estimates may have been influenced by sample attrition over time. Strengths of the study include the cross-national sample that enables us to demonstrate overall consistency of findings across the two geographical locations; the investigation of predictors of mental health of children, which is less common in the COVID-19 literature,^49,50^ and the use of sophisticated analytic techniques that allow us to gauge the relative importance of multiple predictors on mood symptoms.

As vaccination rates increase and society returns to “normal,” the mental health needs of those with continued psychological distress will need to be addressed.^51^ Longitudinal studies are vital for understanding who is going to continue to struggle post-pandemic. This study suggests that in addition to well-established risk factors for post-disaster mental health, attending to the heterogeneity in the impact of the pandemic on people’s daily patterns of interaction and living may provide useful targets for interventions to reduce mood and anxiety symptom levels.

## Data Availability

Data has been made publicly available for download here: http://www.crisissurvey.org/

## Acknowledgements

We thank the CRISIS AFAR team for suggestions to incorporate the OxCGRT into our analyses. Aki Nikolaidis is funded by a NARSAD Young Investigator grant and NIMH grant R21 MH118556-01. This study was funded in part by the financial firm Morgan Stanley. Drs. Paksarian and Merikangas are supported by the Intramural Research Program of the National Institute of Mental Health (ZIA MH002953).

