## Supplemental Material for "Heterogeneity in COVID-19 Pandemic-Induced Lifestyle Stressors and Predicts Future Mental Health in Adults and Children in the US and UK"

**Supplemental Materials**

**Supplemental Methods**

Sample Description- Compared to those who dropped out after April 2020, adults retained in the November analytic sample were older, more commonly white, more likely to be working and less likely to be in school in April, and had lower Prior and Current Mood scores, while children retained in the November parent-report sample endorsed less financial difficulty in April (eTable 2). Compared to population data from the US and UK (in 2018 and 2011 respectively), our adult November samples were younger and more commonly female. While employment status and the proportion of white/non-white persons were similar, the US samples had fewer black and more Asian participants compared to the population. Perceived physical and mental health were worse among US adults but better among children compared to the US population average (eTable 3).

Psychometric Analysis. We used confirmatory factor analysis (CFA) to assess the psychometric structure of the prior mood, current mood, and COVID-19 worries domains across the three timepoints collected (April, May, November). We assessed the quality of unidimensional fit for all items loading onto a single factor using confirmatory fit index (CFI), Tucker-Lewis Index (TLI), root mean square error of approximation (RMSEA), chi-squared, and Omega reliability, and these results are reported in eTable 4.

Life Change Stress Clustering analysis. We use bagging-enhanced Louvain Community Detection to discover groups of individuals that have similar profiles across the life change stress questions. Louvain Community Detection is a clustering approach that finds robust subtypes of individuals through an iterative modularity-optimizing procedure. Most clustering approaches (K-means, spectral clustering), require the experimenter to choose the resolution of the clustering a priori, which can be problematic and lead to instability across samples. Louvain Community detection on the other hand, chooses the cluster resolution that maximizes the modularity of the network. We enhance the reproducibility of our subtyping method through the use of bootstrap aggregation, or bagging. Using bootstrap aggregated clustering creates more reproducible clusters by reducing variability that may occur due to random variations in sample composition.

Predicting Future Mood. Random Forest (RF) creates many single that are comprised of a random selection of variables and a bootstrapped sample is used to train each decision tree. For each iteration of the 1000 bootstrap runs, the performance on each of these decision trees on the out-of-sample data, roughly ⅓ of the sample, is aggregated and used to assess the performance of the RF model. RF provides a robust assessment of the relative impact of each of these variables in predicting outcomes, known as variable importance, which we assess for each variable in our predictive model. Random forest variable importance is known to be biased towards continuous variables (1-4). To address this issue we use the Conditional Random Forest Party package in R, which provides an unbiased tree algorithm with which random forest is calculated. The variable importance mesure from this method is unbiased towards binary vs continuous variables, and is also able to assess the relative importance of each variable given all other variables included in the model.

1) H. Kim and W. Loh. Classification trees with unbiased multiway splits. Journal of the American Statistical Association, 96(454):589–604, 2001.
2) C. Strobl, A.-L. Boulesteix, and T. Augustin. Unbiased split selection for classification trees based on the Gini index. Computational Statistics & Data Analysis, 52(1):483–501, 2007a.
3) C. Strobl, A.-L. Boulesteix, A. Zeileis, and T. Hothorn. Bias in random forest variable importance measures: Illustrations, sources and a solution. BMC Bioinformatics, 8:25, 2007b.
4) T. Hothorn, K. Hornik, and A. Zeileis. Unbiased recursive partitioning: A conditional inference framework. Journal of Computational and Graphical Statistics, 15(3):651–674, 2006.

**Supplemental Results**

Psychometric Structure. We confirmed the unidimensional structure of these domains through CFA across samples and timepoints in these longitudinal data using the "compareFit" function in R (https://cran.r-project.org/web/packages/semTools/citation.html). We used a comparative fit index (CFI) of > 0.90 and an Omega of > 0.8 indicating adequate fit (eTable 4). All time point models were compared against the other time points for adults and children. The chi-square difference test was not significant for any of the comparisons, indicating the unidimensional models fit the data at each timepoint equally well.

Life change stress profiles. Life change stress subtype profiles were highly consistent in structure across April, May and November  (eFigure 1; Pearson’s correlations: adults, April-May: r=0.87-0.99, May-November: r=0.90-0.97; Children, April-May: r=0.96-0.99, May-November: r=0.90-0.95). We found significant differences across LC Subtypes in COVID Worries for April, May, and November, with subtype 2- the socioemotional stress subtype showing the worst scores in each domain (eTable 8). Current Mood and Prior Mood also showed significantly worse scores for the second subtype compared to the other two subtypes, except for April and November Prior Mood, where subtype 2 and 3 had similar scores. The parent report data wholly confirmed this pattern of findings, with the socioemotional financial stress subtype showing significantly worse COVID Worries, Current Mood, and Prior Mood scores than the low stress subtype (1), and the moderate stress subtype (3). For both adult and parent report, the moderate stress subtype also tended to show significantly worse COVID Worries, Prior Mood, and Current Mood state scores compared to the low stress subtype.

Demographics of Life change stress subtypes. Correlates of May life change stress subtypes are shown in eTable 8. COVID worries, prior mood, and current mood all differed by subtype (all p<.001) and were highest in the Higher Social/Higher Economic stress subtype and lowest in the Lower Social/Lower Economic stress subtype, for adults and children. Adults over age 50 were more likely to be in the Lower Social/Lower Economic stress subtype (35.7% versus 19.1% [Higher Social/Higher Economic] and 26.2% [Lower Social/Higher Economic]), while adults under 30 were less likely to be in that subtype (23.0% versus 40.0% [Higher Social/Higher Economic] and 38.2% [Lower Social/Higher Economic]). Children aged 14-17 were more likely to be in the Lower Social/Lower Economic rather than the Higher Social/Higher Economic subtype (21.9% vs 13.3%, p<0.001). Among adults, white participants were more likely to be in the Lower Social/Lower Economic subtype and less likely to be in the Higher Social/Higher Economic stress subtype (p<0.05). Members of the Lower Social/Lower Economic stress subtype were most likely to be working in May (54.6% versus 45.5% [Higher Social/Higher Economic] and 34.9% [Lower Social/Higher Economic]; p < 0.001).

**eFigure 1: Life Change Stress Subtypes by Time Point**

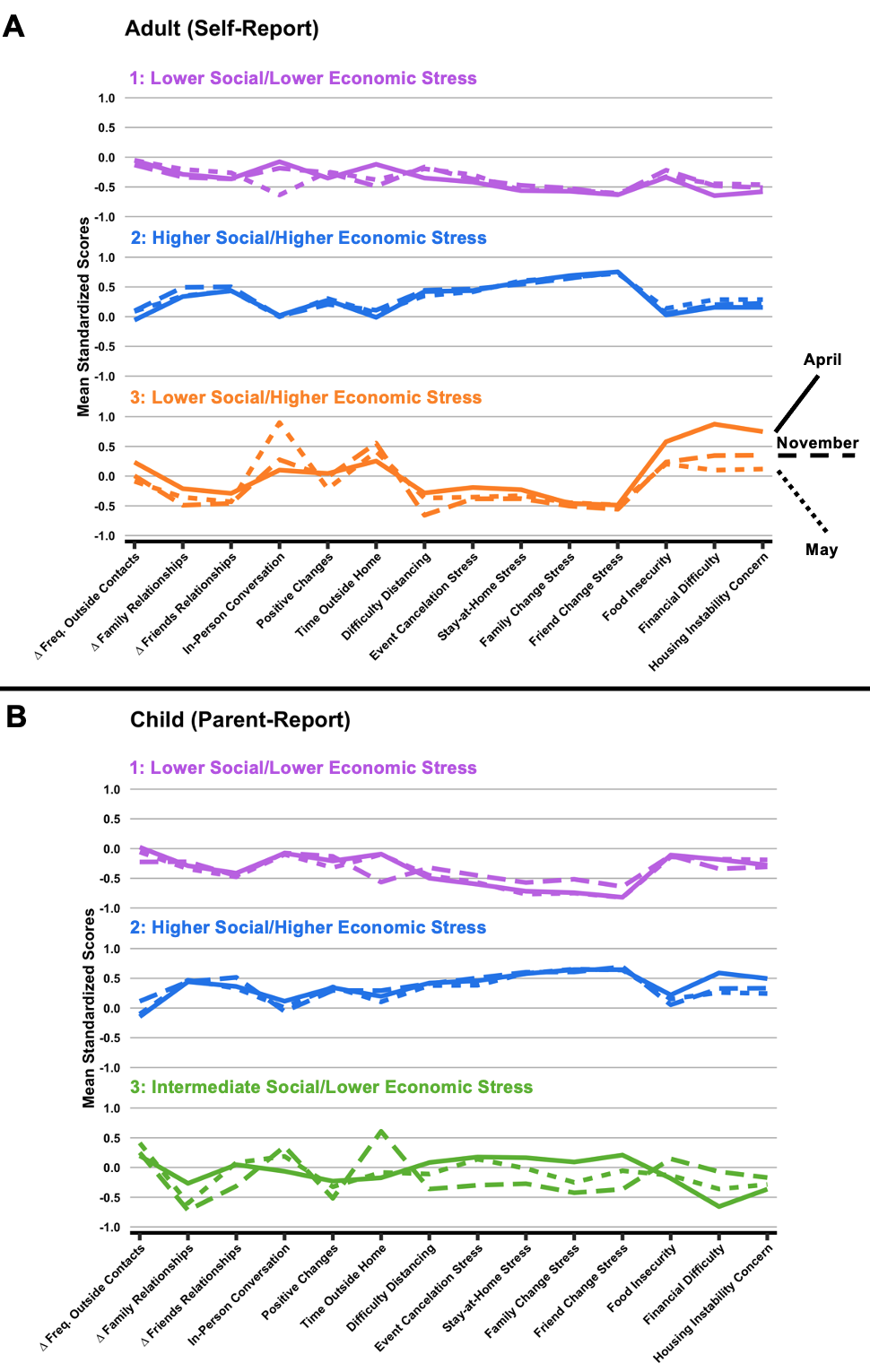

A. Life Change Stress Subtype profiles for adults (top) and children (bottom) across April (straight line), May (short dash), and November (long dash) 2020. Mean normalized profile loadings are displayed on the y-axis. ∆ Family Relationships and ∆ Friends Relationships are coded so that higher scores indicate worsening quality of relationships. In-Person Conversation, Positive Changes, and Time Outside Home are coded so that higher scores indicate less conversations, changes, and time spent outside. Adult Report: Purple (1): Lower Social/Lower Economic Stress, Blue (2): Higher Social/Higher Economic Stress, Orange (3): Lower Social/Higher Economic Stress. Parent Report: Purple (1): Lower Social/Lower Economic Stress, Blue (2): Higher Social/Higher Economic Stress, Orange (3): Intermediate Social/Lower Economic Stress.

**eFigure 2: Predicting November Mood with RF using COVID-19 Threat Data**

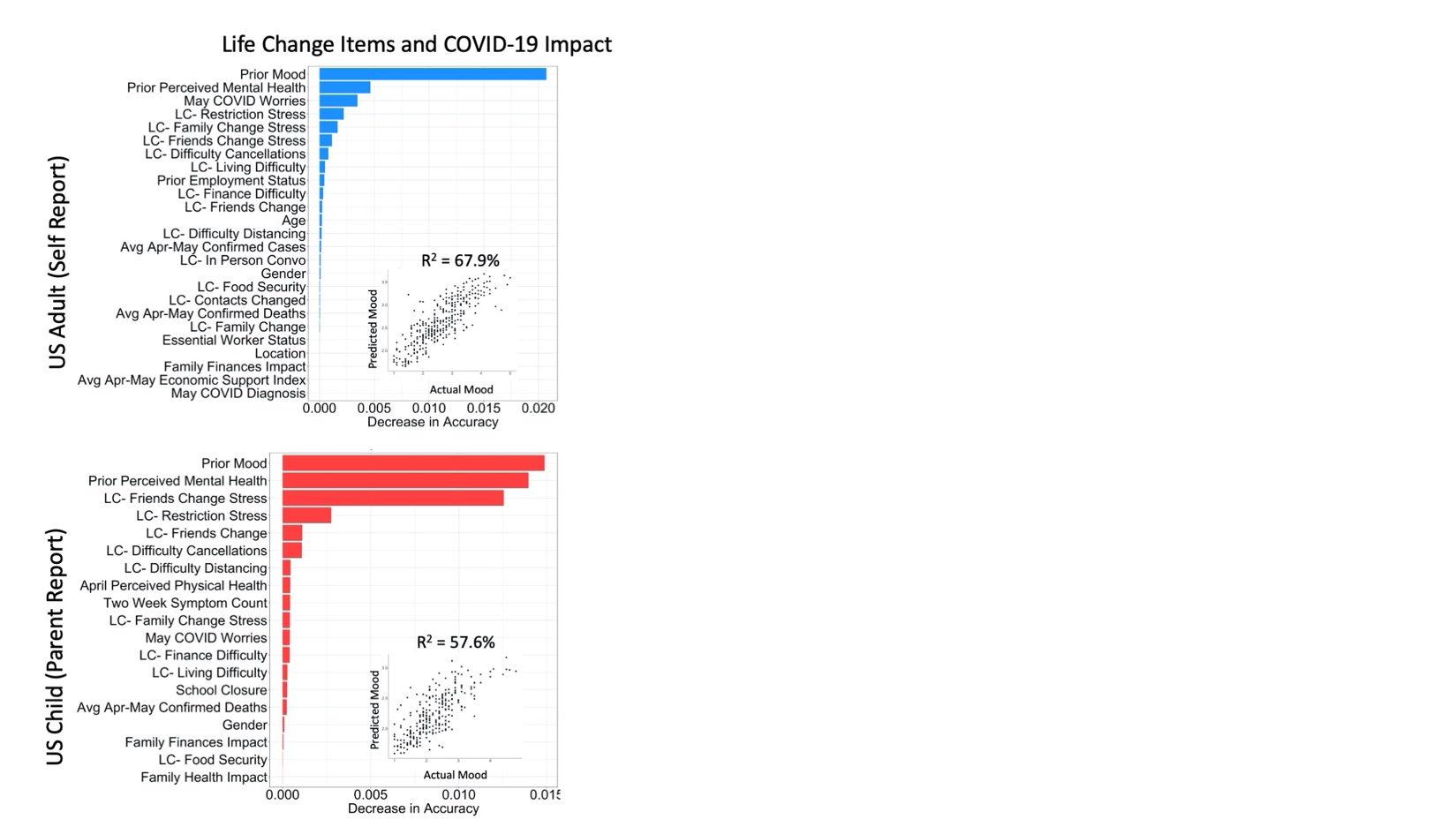

**eTable 1: Analytic Sample Sizes Across Timepoints**

|  | Time point | | |
| --- | --- | --- | --- |
|  | April | May | November |
| Total Adults | 1793 | 1380 | 859 |
| US Adult  California  New York | 895  223  231 | 653  165  168 | 354  79  91 |
| UK Adults  London  Manchester | 898  190  147 | 727  149  121 | 505  105  83 |
| Total Parents | 1466 | 1173 | 780 |
| US Parents  California  New York | 631  71  28 | 459  50  21 | 289  23  14 |
| UK Parents  London  Manchester | 835  120  131 | 714  94  102 | 491  61  62 |

Note: Summary fo the sample sizes with complete data for each timepoint and country.

**eTable 2: Characteristics of the November sample compared to those lost to follow-up**

|  | Adult (Self Report) | | | | | Child (Parent Report) | | | | |
| --- | --- | --- | --- | --- | --- | --- | --- | --- | --- | --- |
|  | Retained  *n* = 859 | | Lost to follow-up  *n* = 934 | |  | Retained  *n* = 780 | | Lost to follow-up  *n* = 686 | |  |
|  | *n or M (SD)* | % | *n or M (SD)* | % | Post-hoc | *n or M (SD)* | % | *n or M (SD)* | % | Post-hoc |
| Age in April | * * * | | | |  |  |  |  |  |  |
| Under 30  30-49  50 and Over | 230  351  278 | 26.8%  40.9%  32.4% | 426  353  155 | 45.6%  37.8%  16.6% | <.001  NS  <.001 |  |  |  |  |  |
| Child Age in April |  |  |  |  |  | – | | | |  |
| 5 and Under  6-13  14-17  18 and Over |  |  |  |  |  | 132  470  164  14 | 16.9%  60.3%  21.0%  1.8% | 108  408  146  24 | 15.7%  59.5%  21.3%  3.5% |  |
| Race/ethnicity | * * * | | | |  | – | | | |  |
| Asian  Black  Hispanic  Other  White | 70  51  58  35  645 | 8.1%  5.9%  6.8%  4.1%  75.1% | 117  60  102  55  600 | 12.5%  6.4%  10.9%  5.9%  64.2% | <.05  NS  <.01  NS  <.001 | 30  40  46  28  636 | 3.8%  5.1%  5.9%  3.6%  81.5% | 20  32  59  25  550 | 2.9%  4.7%  8.6%  3.6%  80.2% |  |
| Gender | – | | | |  | – | | | |  |
| Male  Female  Other | 360  495  4 | 41.9%  57.6%  0.5% | 407  522  5 | 43.6%  55.9%  0.5% |  | 418  360  2 | 53.6%  46.2%  0.3% | 356  327  3 | 51.9%  47.7%  0.4% |  |
| April Job Loss | * * | | | |  |  |  |  |  |  |
| Job Prior, Still Working  Job Prior, Not Still Working  Did Not Have a Job Prior | 409  190  251 | 48.1%  22.4%  29.5% | 419  264  238 | 45.5%  28.7%  25.8% | <.01  NS  NS |  |  |  |  |  |
| April School Closure | * * * | | | |  | – | | | |  |
| School Closed  School Did Not Close  Not Applicable | 117  19  708 | 13.9%  2.3%  83.9% | 226  25  668 | 24.6%  2.7%  72.7% | <.001  NS  <.001 | 710  29  39 | 91.3%  3.7%  5.0% | 616  29  38 | 90.2%  4.2%  5.6% |  |
| April Financial Difficulty | * | | | |  | * * | | | |  |
| Not at All  Slightly  Moderately  Very  Extremely | 268  280  173  91  47 | 31.2%  32.6%  20.1%  10.6%  5.5% | 239  293  227  113  62 | 25.6%  31.4%  24.3%  12.1%  6.6% | NS  NS  NS  NS  NS | 292  246  148  61  33 | 37.4%  31.5%  19.0%  7.8%  4.2% | 235  193  128  87  43 | 34.3%  28.1%  18.7%  12.7%  6.3% | NS  NS  NS  <.05  NS |
| April LC Subtype | * | | | |  | – | | | |  |
| 1- Low Stress  2- Social/High Stress  3- Financial/Moderate | 343  364  152 | 39.9%  42.4%  17.7% | 318  418  198 | 34.0%  44.8%  21.2% | NS  NS  NS | 295  281  204 | 37.8%  36.0%  26.2% | 244  287  155 | 35.6%  41.8%  22.6% |  |
| April COVID Worries | 3.1 (0.7) | – | 3.1 (0.7) |  |  | 2.3 (0.7) | – | 2.4 (0.7) |  |  |
| April Prior Mood | 2.2 (0.7) | * * * | 2.4 (0.7) |  |  | 1.9 (0.6) | – | 2.0 (0.6) |  |  |
| April Current Mood | 2.8 (0.8) | * * | 2.9 (0.8) |  |  | 2.3 (0.7) | – | 2.4 (0.7) |  |  |

*Note.* Participants followed-up in November 2020 are compared to those lost to follow-up. Chi-square and one way analysis of variance tests were used to explore differences across retained and dropout groups for the adult sample and for the parent sample. Post-hoc analyses were conducted as appropriate. * = *p*<0.05, ** = *p*<0.01, *** = *p*<0.001.

**eTable 3: Comparison of analytic sample to US and UK population data**

|  | US | | | UK | | |
| --- | --- | --- | --- | --- | --- | --- |
|  | US 2018 Population | Adult (Self Report) | Child (Parent Report) | UK 2011 Population | Adult (Self Report) | Child (Parent Report) |
| Child Age  0-4  5-17 | 26.73%  73.27% |  | 2.77%  97.23% | 29.27%  70.73% |  | 5.64%  94.36% |
| Age  18-44  45-64  65+ | 45.52%  32.85%  21.63% | 72.07%  23.58%  4.36% |  | 46.95%  32.27%  20.78% | 67.15%  27.28%  5.57% |  |
| Sex  Female  Male | 51.05%  48.95% | 57.38%  42.62% | 53.66%  46.34% | 50.82%  49.18% | 56.63%  43.37% | –  52.46%  47.43% |
| Race/ethnicity ^a^  Asian  Black  Hispanic  Other  White | 6.17%  12.29%  18.46%  3.40%  59.68% | 13.74%  7.93%  15.98%  5.47%  56.87% | 2.54%  5.55%  14.42%  4.44%  73.06% | 7.82%  3.48%  –  3.28%  85.42% | 7.13%  4.45%  –  6.46%  81.96% | 4.07%  4.43%  –  4.67%  86.83% |
| Employment ^b^  Employed  Not Employed | 49.37%  50.63% | 70.84%  28.16% |  | 62.10%  37.90% | 73.22%  26.78% |  |
| Physical Health ^c^  Excellent  Very Good  Good  Fair  Poor | 30.45%  33.29%  26.00%  7.86%  2.40% | 13.65%  33.67%  36.80%  13.53%  2.35% | 49.92%  40.67%  8.13%  1.12%  0.16% | 47.17%  34.22%  13.12%  4.25%  1.25% | 11.40%  37.09%  34.75%  13.07%  3.69% | 49.10%  36.25%  11.16%  2.64%  0.84% |
| Mental Health  Excellent  Very Good  Good  Fair  Poor | 37.87%  30.95%  23.87%  5.81%  1.50% | 17.45%  28.52%  33.45%  15.66%  4.92% | 41.75%  37.30%  15.24%  5.08%  0.63% |  |  |  |

Note: The November US sample is compared to data from the 2018 Medical Expenditures Panel Survey ( <https://www.meps.ahrq.gov/mepstrends/hc_use/>). All variables were measured in April 2020. The November UK sample is compared to data from the last decennial UK census (March 2011), downloaded from: <https://www.nomisweb.co.uk/census/2011>.

^a^ In the child (parent report) sample, parent race was used as a proxy for child race, and was combined with parent-reported information on child’s Hispanic ethnicity.

^b^ Sample employment is based on reports of pre-pandemic employment.

^c^ Levels given below were used in the analytic sample and the US census. Physical health in the UK census was rated as “very good health”, “good health”, “fair health”, “bad health” and “very bad health.”

**eTable 4: CFA Fit for mood and COVID Worries constructs by sample**

| **Factor** | **Sample** | **CFI** | **TLI** | **RMSEA** | **𝜒2** | **Omega (ω)** |
| --- | --- | --- | --- | --- | --- | --- |
| Prior Mood | Adults April | 0.956 | 0.943 | 0.079 | *** | 0.907 |
|  | Adults November | 0.966 | 0.957 | 0.071 | *** | 0.908 |
|  | Children April | 0.926 | 0.905 | 0.091 | *** | 0.884 |
|  | Children November | 0.952 | 0.938 | 0.077 | *** | 0.894 |
| Current Mood | Adults April | 0.96 | 0.949 | 0.077 | *** | 0.911 |
|  | Adults May | 0.965 | 0.955 | 0.069 | *** | 0.903 |
|  | Adults November | 0.978 | 0.972 | 0.064 | *** | 0.914 |
|  | Children April | 0.947 | 0.932 | 0.081 | *** | 0.899 |
|  | Children May | 0.953 | 0.94 | 0.071 | *** | 0.881 |
|  | Children November | 0.95 | 0.936 | 0.087 | *** | 0.9 |
| COVID Worries | Adults April | 0.992 | 0.987 | 0.05 | *** | 0.855 |
|  | Adults May | 0.988 | 0.981 | 0.068 | *** | 0.816 |
|  | Adults November | 0.979 | 0.964 | 0.099 | *** | 0.84 |
|  | Children April | 0.986 | 0.977 | 0.079 | *** | 0.865 |
|  | Children May | 0.984 | 0.974 | 0.079 | *** | 0.878 |
|  | Children November | 0.985 | 0.975 | 0.098 | *** | 0.864 |

Note: Chi-square (𝜒2**)** p values are represented by asterisks, * p < 0.05, ** p < 0.01, *** p < 0.001.

**eTable 5: Unadjusted Sample characteristics with mean COVID Worries, Prior Mood, and Current Mood**

|  | Adults (Self Report) | | | | Child (Parent Report) | | | |
| --- | --- | --- | --- | --- | --- | --- | --- | --- |
|  | *n (%)* | COVID Worries | Prior Mood | Current Mood | *n (%)* | COVID Worries | Prior Mood | Current Mood |
| Country  US  UK | 355 (41.33%)  504 (58.67%) | 2.71 (0.74)  2.72 (0.76) | 2.19 (0.67)  2.19 (0.70) | 2.65 (0.83)  2.63 (0.77) | 489 (62.77%)  290 (37.23%) | 2.14 (0.65)  2.19 (0.69) | 1.84 (0.62)  1.85 (0.61) | 2.21 (0.72)  2.14 (0.66) |
| Age  Under 30  30-49  50 and Over | 213 (28.06%)  267 (35.18%)  279 (36.76%) | 2.66 (0.77)  2.77 (0.76)  2.68 (0.71) | 2.41 (0.72)  2.21 (0.66)  1.99 (0.64) | 2.83 (0.79)  2.72 (0.78)  2.39 (0.77) |  |  |  |  |
| Child Age  5 and Under  6-13  14-17  18 and Over |  |  |  |  | 131(22.59%)  302 (52.07%)  133 (22.93%)  14 (2.41%) | 1.91 (0.6)  2.21 (0.68)  2.28 (0.66)  2.27 (0.46) | 1.69 (0.5)  1.85 (0.62)  1.97 (0.67)  2.22 (0.7) | 2.02 (0.66)  2.2 (0.68)  2.21 (0.69)  2.16 (0.63) |
| Sex  Female  Male  Non-binary  Other  Transmale | 491 (57.16%)  358 (41.68%)  7 (0.81%)  2 (0.23%)  1 (0.12%) | 2.8 (0.74)  2.59 (0.74)  3.05 (0.74)  3.17 (0.47)  3.17 (NA) | 2.26 (0.68)  2.07 (0.69)  2.53 (0.33)  2.55 (0.07)  2.5 (NA) | 2.76 (0.79)  2.47 (0.78)  3.24 (0.4)  2.35 (1.48)  3.4 (NA) | 362 (46.41%)  414 (53.08%)  2 (0.26%)  0 (0.00%)  2 (0.26%) | 2.18 (0.66)  2.17 (0.69)  2.11 (0.69)  NA  2.08 (0.35) | 1.79 (0.61)  1.9 (0.63)  1.73 (0.15)  NA  2.3 (0.57) | 2.16 (0.7)  2.17 (0.68)  2.17 (0.42)  NA  2.5 (0.14) |
| Race  Asian  Black  Hispanic  White  Other | 70 (8.15%)  51(5.94%)  58 (6.75%)  645 (75.09%)  35 (4.07%) | 2.64 (0.69)  2.7 (0.74)  2.65 (0.85)  2.76 (0.77)  2.73 (0.74) | 2.23 (0.59)  2.28 (0.73)  2.18 (0.78)  2.43 (0.67)  2.16 (0.69) | 2.64 (0.75)  2.69 (0.73)  2.68 (0.96)  2.95 (0.84)  2.62 (0.79) | 30 (3.85%)  40 (5.13%)  46 (5.90%)  636 (81.54%)  28 (3.59%) | 1.87 (0.46)  2.22 (0.64)  2.22 (0.67)  1.96 (0.79)  2.19 (0.68) | 1.71 (0.5)  1.76 (0.7)  2.09 (0.77)  1.9 (0.68)  1.84 (0.6) | 2.02 (0.5)  1.79 (0.63)  2.5 (0.76)  2.12 (0.69)  2.18 (0.68) |
| Urbanicity  Large City  Suburbs of a Large City  Small City  Town or Village  Rural Area | 198 (23.05%)  237 (27.59%)  128 (14.90%)  241 (28.06%)  55 (6.40%) | 2.77 (0.77)  2.71 (0.77)  2.69 (0.68)  2.70 (0.74)  2.62 (0.77) | 2.22 (0.68)  2.17 (0.67)  2.27 (0.76) 2.14 (0.68)  2.18 (0.69) | 2.74 (0.78)  2.68 (0.79(  2.62 (0.81)  2.57 (0.80)  2.46 (0.80) | 110 (14.12%)  246 (31.58%)  96 (12.32%)  272 (34.92%)  55 (7.06%) | 2.23 (0.72)  2.15 (0.59)  2.23 (0.78)  2.11 (0.67)  2.36 (0.64) | 1.82 (0.66)  1.80 (0.59)  1.93 (0.66)  1.84 (0.62)  1.95 (0.57) | 2.21 (0.80)  2.18 (0.71)  2.19 (0.63)  2.12 (0.64)  2.28 (0.65) |
| Education  Some Grade School  Some High School  High School Diploma or GED  Some College or 2-Year Degree  4-Year College Graduate  Some School Beyond College  Graduate or Professional Degree | 4 (0.47%)  16 (1.87%)  120 (14.00%)  229 (26.72%)  247 (28.82%)  25 (2.92%)  216 (25.20%) | 2.50 (0.00)  2.71 (0.83)  2.77 (0.79)  2.69 (0.73)  2.67 (0.69)  2.58 (0.83)  2.77 (0.78) | 2.40 (0.42)  2.25 (0.68)  2.25 (0.75)  2.25 (0.72)  2.13 (0.71)  2.14 (0.92)  2.14 (0.58) | 2.45 (0.35)  2.65 (0.83)  2.70 (0.84)  2.65 (0.80)  2.55 (0.76)  2.53 (0.94)  2.69 (0.79) | 3 (0.39%)  23 (2.96%)  87 (11.18%)  204 (26.22%)  235 (30.21%)  29 (3.73%)  197 (25.32%) |  |  |  |
| Essential Worker in Family  No  Yes | 646 (75.56%)  209 (24.44%) | 2.68 (0.73)  2.77 (0.78) | 2.15 (0.7)  2.25 (0.68) | 2.61 (0.82)  2.7 (0.76) | 427 (54.95%)  350 (45.05%) | 2.19 (0.66)  2.16 (0.69) | 1.86 (0.65)  1.84 (0.58) | 2.13 (0.65)  2.22 (0.72) |
| Any Family Impact  No  Yes | 500 (58.41%)  356 (41.59%) | 2.62 (0.75)  2.85 (0.73) | 2.11 (0.68)  2.3 (0.68) | 2.54 (0.8)  2.79 (0.77) | 515 (66.28%)  262 (33.72%) | 2.14 (0.68)  2.24 (0.65) | 1.82 (0.62)  1.9 (0.62) | 2.09 (0.63)  2.34 (0.74) |
| Family Member Dx  No  Yes | 739 (86.13%)  119 (13.87%) | 2.69 (0.75)  2.85 (0.72) | 2.17 (0.69)  2.31 (0.70) | 2.62 (0.79)  2.76 (0.81) | 695 (89.79%)  79 (10.21%) | 2.16 (0.67)  2.3 (0.71) | 1.84 (0.61)  1.92 (0.73) | 2.16 (0.68)  2.33 (0.7) |
| COVID Diagnosis  No  Yes | 839 (97.8%)  19 (2.2%) | 2.71 (0.75)  2.96 (0.67) | 2.18 (0.69)  2.50 (0.68) | 2.64 (0.80)  2.89 (0.77) | 769 (98.7%)  10 (1.3%) | 2.17 (0.67)  2.67 (0.81) | 1.84 (0.61)  2.56 (0.92) | 2.16 (0.68)  2.74 (0.87) |
| 2-Week Symptom Count  None  One  Two  Three or More | 411 (47.85%)  138 (16.07%)  100 (11.64%)  210 (24.45%) | 2.58 (0.74)  2.63 (0.64)  2.92 (0.71)  2.93 (0.78) | 2.06 (0.69)  2.16 (0.65)  2.25 (0.62)  2.42 (0.69) | 2.45 (0.78)  2.6 (0.77)  2.81 (0.74)  2.95 (0.76) | 431 (55.26%)  119 (15.26%)  109 (13.97%)  121 (15.51%) | 2.13 (0.65)  2.18 (0.64)  2.18 (0.71)  2.34 (0.75) | 1.8 (0.61)  1.92 (0.6)  1.8 (0.57)  2.02 (0.67) | 2.1 (0.68)  2.16 (0.64)  2.17 (0.7)  2.43 (0.67) |

*Note:* N=859 adults and N=780 children assessed via parent report. Unadjusted mean scores and standard deviations of COVID Worries, Prior Mood, and Current Mood (November) presented across key demographic variables.

**eTable 6: Adjusted associations between sample characteristics and COVID Worries, Current Mood and Prior Mood**

|  | Adult (Self Report) - November | | | Child (Parent Report) - November | | |
| --- | --- | --- | --- | --- | --- | --- |
|  | COVID Worries | April Prior Mood | Current Mood | COVID Worries | April Prior Mood | Current Mood |
| Country   1. US 2. UK | 1) Reference  2) 0.0 | 1) Reference  2) 0.015 | 1) Reference  2) -0.034 | 1) -0.09  2) Reference | 1) 0.03  2) Reference | 1) 0.05  2) Reference |
| Age   1. Under 30 2. 30-49 3. Over 50 | 1) -0.14*  2) Reference  3) -0.004 | 1) 0.14*  2) Reference  3) -0.21*** | 1) 0.047  2) Reference  3) -0.27 |  |  |  |
| Child Age   1. 5 and Under 2. 6-13 3. 14-17 4. 18 and Over |  |  |  | 1) -0.15*  2) Reference  3) -0.002  4) 0.07 | 1) -0.16**  2) Reference  3) 0.04  4) -0.10 | 1) -0.05  2) Reference  3) -0.02  4) -0.28 |
| Sex   1. Female 2. Male | 1) Reference  2) -0.18** | 1) Reference  2) -0.10* | 1) Reference  2) -0.18 | 1) 0.04  2) Reference | 1) -0.03  2) Reference | 1) 0.03  2) Reference |
| Race   1. Asian 2. Black 3. Hispanic 4. Other 5. White | 1) 0.05  2) -0.17  3) 0.019  4) 0.31*  5) Reference | 1) 0.026  2) 0.013  3) 0.033  4) 0.17  5) Reference | 1) 0.09  2) 0.010  3) 0.057  4) 0.15  5) Reference | 1) -0.39**  2) -0.05  3) 0.02  4) -0.24  5) Reference | 1) -0.12  2) -0.16  3) 0.20*  4) -0.006  5) Reference | 1) -0.21  2) -0.45***  3) 0.28**  4) -0.03  5) Reference |
| Urbanicity   1. Large City 2. Rural 3. Small City 4. Suburbs of Large City 5. Town or Village | 1) 0.01  2) Reference  3) -0.06  4) 0.0  5) -0.10 | 1) 0.002  2) Reference  3) 0.089  4) 0.037  5) 0.077 | 1) -0.01  2) Reference  3) -0.09  4) -0.001  5) -0.15 | 1) 0.18*  2) 0.11  3) 0.12  4) Reference  5) 0.29** | 1) -0.02  2) -0.07  3) -0.008  4) Reference  5) 0.01 | 1) 0.15  2) 0.08  3) 0.05  4) Reference  5) 0.15 |
| Education   1. Some High School 2. High School Diploma or GED 3. Some College or 2-Year Degree* 4. 4-Year College Graduate 5. Some School Beyond College 6. Graduate or Professional Degree | 1) 0.15  2) 0.14  3) Reference  4) 0.01  5) -0.12  6) 0.07 | 1) 0.12  2) 0.02  3) Reference  4) -0.08  5) -0.17  6) -0.11 | 1) 0.18  2) 0.12  3) Reference  4) -0.06  5) -0.15  6) 0.05 |  |  |  |
| Essential Worker in Family   1. Yes 2. No | 1) 0.09  2) Reference | 1) 0.005  2) Reference | 1) 0.024  2) Reference | 1) -0.04  2) Reference | 1) 0.007  2) Reference | 1) 0.07  2) Reference |
| Any Family Impact   1. Yes 2. No | 1) 0.15 **  2) Reference | 1) 0.076  2) Reference | 1) 0.13*  2) Reference | 1) 0.03  2) Reference | 1) 0.05  2) Reference | 1) 0.19***  2) Reference |
| Family COVID Diagnosis   1. Yes 2. No | 1) 0.027  2) Reference | 1) -0.25  2) Reference | 1) 0.029  2) Reference | 1) 0.09  2) Reference | 1) -0.01  2) Reference | 1) -0.02  2) Reference |
| COVID Diagnosis   1. Yes 2. No | 1) 0.16  2) Reference | 1) 0.18  2) Reference | 1) 0.16  2) Reference | 1) 0.35  2) Reference | 1) 0.73***  2) Reference | 1) 0.37  2) Reference |
| Two Week Symptom Count   1. None 2. One 3. Two 4. Three or more | 1) Reference  2) 0.01  3) 0.29 ***  4) 0.26 *** | 1) Reference  2) 0.046  3) 0.13  4) 0.28*** | 1) Reference  2) 0.074  3) 0.25**  4) 0.38*** | 1) Reference  2) 0.03  3) 0.04  4) 0.20** | 1) Reference  2) -0.03  3) 0.12  4) 0.16* | 1) Reference  2) 0.04  3) 0.05  4) 0.27*** |

Note: Adjusted associations were estimated via multiple linear regression. * = p<.05, ** = p<.01, *** = p<.001.All demographic variables are from April 2020 while all COVID Impact variables were measured in November 2020.

**eTable 7: May Life Change Subtype Comparison**

|  |  | Adult (Self Report) | | | | Child (Parent Report) | | | |
| --- | --- | --- | --- | --- | --- | --- | --- | --- | --- |
|  |  | Lower Social/Lower Economic Stress  (1)  *n* =457 | Higher Social/Higher Economic Stress  (2)  *n* = 598 | Lower Social/ Higher Economic Stress  (3)  *n* = 325 |  | Lower Social/Lower Economic Stress  (1)  *n* =420 | Higher Social/Higher Economic Stress  (2)  *n* = 557 | Intermediate Social/ Lower Economic Stress  (3)  *n* = 196 |  |
|  |  | M (SD) or % | M (SD) or % | M (SD) or % | Post-hoc | M (SD) or % | M (SD) or % | M (SD) or % | Post-hoc |
| Life Changes | Change of frequency of outside contacts  M (SD)  Change of family relationships  M (SD)  Change of friend relationships  M (SD)  In-person conversations  M (SD)  Positive changes  M (SD)  Time outside home  M (SD)  Difficulty distancing  M (SD)  Event cancellation stress  M (SD)  Stay-at-home stress  M (SD)  Family change stress  M (SD)  Friend change stress  M (SD)  Food insecurity  M (SD)  Financial insecurity  M (SD)  Housing instability concern  M (SD) | 2.62 (1.4)  3.27 (0.6)  3.06 (0.5)  10.27 (33.1)  1.92 (0.8)  2.06 (1.3)  1.77 (0.9)  2.10 (1.1)  2.02 (0.8)  1.60 (0.8)  1.55 (0.7)  0.07 (0.2)  1.69 (0.9)  1.57 (0.8) | *  2.41 (1.6)  * * *  2.86 (0.8)  * * *  2.61 (0.7)  * * *  4.04 (8.4)  * * *  1.49 (0.7)  * * *  2.51 (1.2)  * * *  2.32 (1.1)  * * *  3.00 (1.2)  * * *  3.26 (1.0)  * * *  2.98 (1.1)  * * *  3.04 (1.0)  * * *  0.25 (0.4)  * * *  2.54 (1.2)  * * *  2.45 (1.3) | 2.68 (1.5)  3.38 (0.7)  3.17 (0.6)  0.91 (3.2)  1.89 (0.8)  2.08 (1.1)  1.59 (0.9)  2.02 (1.2)  2.26 (0.9)  2.71 (0.9)  1.69 (0.8)  0.28 (0.4)  2.33 (1.2)  2.25 (1.2) | 3>2  1=3>2  3>1>2  1>2=3  1=3>2  1>2>3  2>1>3  2>1=3  2>3>1  2>3>1  2>1=3  2=3>1  2>3>1  2>3>1 | 2.42 (1.5)  3.47 (0.8)  2.89 (0.6)  2.64 (4.0)  1.90 (0.8)  2.50 (1.3)  1.56 (0.8)  1.73 (0.9)  1.71 (0.7)  1.41 (0.6)  1.55 (0.7)  0.06 (0.2)  1.88 (1.0)  1.27 (0.6) | * * *  2.50 (1.7)  * * *  2.78 (0.8)  * * *  2.27 (0.8)  –  2.32 (3.2)  * * *  1.38 (0.6)  * *  2.23 (1.3)  * * *  2.45 (1.1)  * * *  2.89 (1.2)  * * *  3.15 (1.0)  * * *  2.95 (1.0)  * * *  3.16 (1.0)  * * *  0.15 (0.4)  * * *  2.37 (1.2)  * * *  1.62 (0.9) | 1.67 (1.3)  3.69 (0.7)  2.46 (0.8)  2.22 (7.5)  1.90 (0.8)  2.47 (1.4)  1.92 (0.9)  2.59 (1.1)  2.51 (0.7)  1.96 (0.8)  2.40 (0.7)  0.06 (0.2)  1.68 (0.9)  1.19 (0.5) | 1=2>3  3>1>2  1>3>2  1=3>2  1>2  2>3>1  2>3>1  2>3>1  2>3>1  2>3>1  2>1=3  2>1=3  2>1=3 |

*Note*. *N*=1,380 adults and *N*=1,173 children assessed via parent report. One-way analysis of variance tests were conducted to test group differences. All variables were assessed at the May time point.

**eTable 8. Adult and Parent Report Subtype ANOVAs**

|  |  |  | Adult LC Subtype | | |  |  |  |
| --- | --- | --- | --- | --- | --- | --- | --- | --- |
| Sample |  | Mood | 1  (Low Stress) | 2  (Socioemotional Stress) | 3  (Financial Stress) | F | Cohen's d | Post Hoc |
| Adult | April | COVID Worries | 2.84 (0.66) | 3.28 (0.71) | 3.15 (0.71) | 76.13*** | 13.67 | 2 > 3 > 1 |
|  |  | Prior Mood | 2.08 (0.65) | 2.41 (0.74) | 2.39 (0.69) | 45.55*** | 12.39 | 2 = 3 > 1 |
|  |  | Current Mood | 2.4 (0.65) | 3.17 (0.75) | 2.88 (0.74) | 206.4*** | 7.24 | 2 > 3 > 1 |
|  | May | COVID Worries | 2.67 (0.64) | 3.16 (0.71) | 2.82 (0.74) | 66.05*** | 11.48 | 2 > 3 > 1 |
|  |  | Current Mood | 2.32 (0.61) | 3.09 (0.71) | 2.53 (0.69) | 179.75*** | 6.65 | 2 > 3 > 1 |
|  | November | COVID Worries | 2.43 (0.67) | 2.95 (0.71) | 2.66 (0.77) | 44.43*** | 10.28 | 2 > 3 > 1 |
|  |  | Prior Mood | 2.01 (0.66) | 2.32 (0.68) | 2.2 (0.7) | 17.53*** | 13.93 | 2 = 3 > 1 |
|  |  | Current Mood | 2.23 (0.66) | 3.01 (0.74) | 2.52 (0.76) | 99.93*** | 6.56 | 2 > 3 > 1 |
|  |  |  | Child LC Subtype | | |  |  |  |
| Sample |  | Mood | 1  (Low Stress) | 2 (Socioemotional Financial Stress) | 3  (Moderate Stress) | F | Cohen’s d | Post Hoc |
| Child | April | COVID Worries | 2.06 (0.58) | 2.61 (0.78) | 2.38 (0.63) | 92.85*** | 8.51 | 2 > 3 > 1 |
|  |  | Prior Mood | 1.79 (0.55) | 2.11 (0.72) | 1.91 (0.57) | 36.59*** | 11.98 | 2 > 3 > 1 |
|  |  | Current Mood | 1.94 (0.52) | 2.77 (0.72) | 2.36 (0.55) | 255.03*** | 5.68 | 2 > 3 > 1 |
|  | May | COVID Worries | 1.98 (0.56) | 2.54 (0.74) | 2.2 (0.59) | 90.2*** | 7.94 | 2 > 3 > 1 |
|  |  | Current Mood | 1.91 (0.46) | 2.68 (0.66) | 2.17 (0.47) | 225.39*** | 5.75 | 2 > 3 > 1 |
|  | November | COVID Worries | 1.92 (0.52) | 2.42 (0.71) | 2.12 (0.66) | 51.35*** | 8.56 | 2 > 3 > 1 |
|  |  | Prior Mood | 1.73 (0.56) | 1.96 (0.63) | 1.84 (0.68) | 11.19*** | 16.02 | 2 > 1 |
|  |  | Current Mood | 1.84 (0.53) | 2.54 (0.66) | 1.93 (0.56) | 123.06*** | 5.52 | 2 > 1 = 3 |

Note: ANOVA tests of subtype differences in COVID Worries, Prior Mood, and Current Mood (November) across all three timepoints. **p*<0.05, ***p*<0.01, ****p*<0.001.

**ADDITIONAL SUPPLEMENTAL TEXT MATERIALS**

Oxford COVID-19 Government Response Tracker

**Confirmed Cases by state population:** number of confirmed COVID-19 cases in the state per 1,000,000 residents.

**Confirmed Deaths by state population:** number of confirmed COVID-19 deaths in the state per 1,000,000 residents.

**Containment and Health Index:** *School Closing, Workplace Closing, Cancel Public Events, Restrictions on Gatherings, Close Public Transport, Stay at Home Requirements, Restrictions on Internal Movement, International Travel Controls, Public Information Campaigns, Testing Policy, Contact Tracing, Facial Coverings, Vaccination Policy, Protection of Elderly People*

**Economic Support Index:** *Income Support, Debt/Contract Relief*

Calculation for indices:

Index = $\frac{1}{k}\sum_{j = 1}^{k} I$*_j_* where k = number of component variables and I_j_ is the sub-index score for each individual variable.

**Life Changes and Mood States**

Life Changes: During the past two weeks...

*School closure during the pandemic*

27. … if you attend school, has your school building been closed? Y/N/Not Applicable

a. If no,
   i. Are classes in session? Y/N
   ii. Are you attending classes in-person? Y/N
b. If yes,
   i. Have classes resumed online? Y/N
   ii. Do you have easy access to the internet and a computer? Y/N
   iii. Are there assignments for you to complete? Y/N
   iv. Are you able to receive meals from the school? Y/N

*Job loss during the pandemic*

28. … if you had a job prior to Coronavirus/COVID-19, are you still working? Y/N/Not Applicable

a. If yes,
   i. Are you still going to your workplace? Y/N
   ii. Are you teleworking or working from home? Y/N
b. If no,
   i. Were you laid off from your job? Y/N
   ii. Do you lose your job? Y/N

*In-person conversation*

29. … how many people, from outside of your household, have you had an in-person conversation with? ____

*Time outside home*

30. … how much time have you spent going outside of the home (e.g., going to stores, parks, etc.)? Scaled 1 (Not at all) to 5 (Every day)

*Stay-at-home stress*

31. … how stressful have the restrictions on leaving home been for you? Scaled 1 (Not at all) to 5 (Extremely)

*Change of frequency of outside contacts*

32. … have your contacts with people outside of your home changed relative to *before* the Coronavirus/COVID-19 crisis in your area? Scaled a (A lot less) to e (A lot more)

*Difficulty distancing*

33. … how much difﬁculty have you had following the recommendations for keeping away from close contact with people? Scaled 1 (None) to 5 (A great amount)

*Change of family relationships*

34. … has the quality of the relationships between you and members of your family changed? Scaled a (A lot worse) to e (A lot better)

*Family change stress*

35. … how stressful have these changes in family contacts been for you? Scaled 1 (Not at all) to 5 (Extremely)

*Change of friends relationships*

36. … has the quality of your relationships with your friends changed? Scaled a (A lot worse) to e (A lot better)

*Friend change stress*

37. … how stressful have these changes in social contacts been for you? Scaled 1 (Not at all) to 5 (Extremely)

*Event cancelation stress*

38. … how much has cancellation of important events (such as graduation, prom, vacation, etc.) in your life been difficult for you? Scaled 1 (Not at all) to 5 (Extremely)

*Financial insecurity*

39. … to what degree have changes related to the Coronavirus/COVID-19 crisis in your area created financial problems for you or your family? Scaled 1 (Not at all) to 5 (Extremely)

*Housing instability concern*

40. … to what degree are you concerned about the stability of your living situation? Scaled 1 (Not at all) to 5 (Extremely)

*Food insecurity*

41. … did you worry whether your food would run out because of a lack of money? 1 (Yes) or 2 (No)

*Positive changes*

42. How hopeful are you that the Coronavirus/COVID-19 crisis in your area will end soon? Scaled 1 (Not at all) to 5 (Extremely)

Mood States: During the past two weeks...

*Worry*

34. … how worried were you generally? Scaled a (Not worried at all) to e (Extremely worried)

*Happy vs. Sad*

35. … how happy versus sad were you? Scaled a (Very sad/depressed/unhappy) to e (Very happy/cheerful)

*Enjoy Activities*

36. … how much were you able to enjoy your usual activities? Scaled a (Not at all) to e (A lot)

*Relaxed vs. Anxious*

37. … how relaxed versus anxious were you? Scaled a (Very relaxed/calm) to e (Very nervous/anxious)

*Fidget*

38. … how fidgety or restless were you? Scaled a (Not fidgety/restless at all) to e (Extremely fidgety/restless)

*Fatigue*

39. … how fatigued or tired were you? Scaled a (Not fatigued or tired at all) to e (Extremely fatigued or tired)

*Focus*

40. … how well were you able to concentrate or focus? Scaled a (Very focused/attentive) to e (Very unfocused/distracted)

*Irritability*

41. … how irritable or easily angered were you? Scaled a (Not irritable or easily angered at all) to e (Extremely irritable or easily angered)

*Loneliness*

42. … how lonely were you? Scaled 1 (Not lonely at all) to 5 (Extremely lonely)

*Negative Thoughts*

43. … to what extent did you have negative thoughts, thoughts about unpleasant experiences or things that made you feel bad? Scaled 1 (Not at all) to 5 (A lot of the time)
